# Comparing the effect profile of CETP in individuals of East Asian and European ancestries

**DOI:** 10.1101/2023.05.26.23290616

**Authors:** Diana Dunca, Sandesh Chopade, María Gordillo-Marañón, Aroon D. Hingorani, Karoline Kuchenbaecker, Chris Finan, Amand F. Schmidt

## Abstract

**Background:** Cholesteryl ester transfer protein (CETP) is a lipid drug target under development for coronary heart disease (CHD) in both European and East Asian populations. Previous drug target Mendelian randomization (MR) studies conducted in East Asians failed to show a CHD effect, which has been interpreted as lack of effectiveness of CETP inhibition for CHD prevention in this population.

**Objectives:** In this study, we inferred the effect of CETP inhibition in individuals of European and East Asian ancestries using drug target Mendelian randomization.

**Methods:** We leveraged genetic associations of *CETP* variants with major blood lipid fractions for individuals of European (n=1,320,016) and East Asian (n=146,492) ancestries. Colocalization was employed to identify potential cross-ancestry signals of *CETP* variants for plasma concentrations of low-density lipoprotein cholesterol (LDL-C) or high-density lipoprotein cholesterol (HDL-C). Drug target MR was used to estimate ancestry-specific effects of on-target *CETP* inhibition. Differences between ancestries were evaluated using interaction tests, applying a multiplicity corrected alpha of 1.9×10^-3^ based on the 26 considered traits.

**Results:** There was strong support (posterior probability: 1.00) of a shared causal *CETP* variant affecting HDL-C in both populations, which was not observed for LDL-C. Employing drug target MR scaled to a standard deviation increase in HDL-C, we found that lower CETP was associated with lower LDL-C, Lp[a], systolic blood pressure and pulse pressure in both groups, but the effects were more pronounced in European individuals (interaction p-values < 1.9×10^-3^). Lower CETP was protective against CHD, angina, intracerebral haemorrhage and heart failure in both ancestries, for example for CHD in East Asians (OR 0.89, 95%CI 0.84;0.94) compared to Europeans (OR 0.95, 95%CI 0.92;0.99, interaction p-value=0.05).

**Conclusions:** In conclusion, on-target inhibition of CETP is anticipated to decrease cardiovascular disease in individuals of both European and East Asian ancestries.

## Introduction

Cholesteryl ester transfer protein (CETP) plays a crucial role in the reverse cholesterol transport from peripheral tissues to the liver by promoting the exchange of triglycerides (TG) and cholesterol ester between high-density lipoprotein cholesterol (HDL-C) and other apolipoprotein-B rich particles, including low-density lipoprotein cholesterol (LDL-C)^1^. Due to its effects on HDL-C and LDL-C, there have been numerous attempts to develop CETP-inhibitor drugs to reduce coronary heart disease (CHD) risk. The REVEAL^2^ trial showed a protective effect of anacetrapib on cardiovascular disease (CVD), but the drug was not pursued to market for commercial or other reasons^3, 4^. We have shown that previous failures of several CETP-inhibitor drugs in clinical trials can be attributed to the compounds or trial duration rather than a failure of the target^5^.

Drug target Mendelian randomization (MR) analyses conducted in European populations, leveraging genomic variants within and around the *CETP* locus, have consistently indicated that sufficiently potent on-target inhibition of CETP is anticipated to decrease CHD risk^5–7^. However, previous MR studies on CETP in East Asian populations have failed to replicate this protective effect on CHD^8^. Loss-of-function *CETP* variants (D442G and Int14A) found in Japanese individuals are associated with a 35% decrease in CETP concentration, as well as a 10% elevation in HDL-C concentration^9–11^. Therefore, the lack of a protective effect of CETP inhibition inferred from MR analysis of CETP and CHD in East Asian populations is unexpected.

Due to ongoing efforts, the availability of genomic data on East Asian participants is growing. Biobank Japan (BBJ)^12^ has released large sample size analyses (n=179,000) covering 220 clinical phenotypes, and biomarkers. Concomitantly, the Global Lipids Genetics Consortium (GLGC)^13^, a multi-ancestry meta-analysis of 201 studies, has published a genome-wide association study (GWAS) including genetic associations with lipid concentrations from 146,492 East Asian participants.

Given the growing number of genotyped East Asian participants, we aimed to conduct a large sample size drug target MR analysis of the on-target effect of CETP inhibition, exploring potential differences between European and East Asian populations. First, colocalization was employed to determine potential cross-ancestry signals between *CETP* variants for HDL-C and LDL-C. Subsequently, we performed a biomarker weighted drug target MR analysis, scaling the CETP effect by a standard deviation increase in HDL-C concentration. Specifically, we considered effects on 26 clinically relevant traits including 11 biomarker traits, cardiovascular outcomes such as CHD, angina, peripheral artery disease (PAD), stroke, intracerebral & subarachnoid haemorrhage, heart failure (HF), as well as potential safety outcomes: chronic kidney disease (CKD), pneumonia, chronic obstructive pulmonary disease (COPD), type 2 diabetes (T2D), asthma, and glaucoma.

## Methods

### Data sources

Genetic association data on plasma LDL-C, HDL-C and TG concentrations were extracted from GLGC, which released aggregated data (i.e., point estimates and standard errors) for 146,492 East Asian participants and 1,320,016 European participants.

The following outcome data was sourced for the drug target MR analyses to estimate the on-target effects of CETP inhibition **(Supplemental Table 1)**. For individuals of European ancestry, we leveraged data on apolipoprotein (Apo) A1 and B, lipoprotein a (Lp[a]), from 361,194 UK biobank (UKB) participants, systolic blood pressure (SBP), diastolic blood pressure (DBP) and pulse pressure (PP) on 757,601 participants^14^, glucose and HbA1c on 196,991 participants^15^, C-reactive protein (CRP, n= 204,402)^16^, CHD (60,801 case)^17^, any stroke (110,182 cases) and ischemic stroke (86,668 cases)^18^, HF (47,309 cases)^19^, T2D (180,834 cases)^20^, CKD (41,395 cases)^21^, glaucoma (15,655 cases)^22^ and subarachnoid haemorrhage (5,140 cases)^23^. Additional outcome data was sourced from a FinnGen and UKB meta-analyses by Sakaue *et. al.*^12^ on angina (30,025 cases), ventricular arrhythmia (1,018 cases), PAD (7,114 cases), asthma (38,369 cases), intracerebral haemorrhage (1,935 cases), pneumonia (16,887 cases), with COPD (58,559 cases) included from the Global Biobank Meta-analysis Initiative (GBMI)^24^.

The corresponding outcomes in the East Asian participants were accessed through the Pan-ancestry GWAS of the UK Biobank (Pan-UKB)^25^ on Apo-A1 (n=2,325), Apo-B (n=2,553) and Lp[a] (n=2,275). Additional cardiometabolic biomarker data was sourced from Biobank Japan (BBJ)^12^ on SBP, DBP, PP, glucose, HbA1c, CRP for between 71,221 and 145,505 participants **(Supplemental Table 1)**. BBJ provided data on CHD (32,512 cases), angina (14,007 cases), PAD (4,112 cases), ischemic stroke (22,664 cases), subarachnoid (1,203 cases) and intracerebral (1,456 cases) haemorrhage, ventricular arrhythmia (1,673 cases), T2D (45,383 cases), CKD (2,117 cases), glaucoma (8,448 cases), pneumonia (7,423 cases).

Finally, the following outcomes were sourced from the East Asian GBMI release: HF (12,665 cases), COPD (19,044 cases), and any stroke (23,345 cases).

### Cross-ancestry colocalization of the LDL-C and HDL-C *CETP* signals

Due to sampling variability as well as linkage disequilibrium (LD), the most significant variant at a given locus may not reflect the causal variant. Colocalization identifies potential shared causal variants between two traits^26^ while accounting for sampling variability and LD. Due to the larger sample size available in the European GLGC GWAS, rs183130 (16:g.56991363C>T, GRCh37) has been robustly identified as a causal *CETP* variant for both LDL-C and HDL-C. We leveraged coloc^27^ to determine whether this European fine-mapped variant was also causal for LDL-C and HDL-C in East Asian participants. We considered genetic variants within a ±50kb flank of the CETP genomic region and a MAF ≥ 0.01, applying the following posterior probabilities: PP.H1, PP.H2 =10^-4^ to detect if at least a single genetic variant was associated with the plasma lipids in Europeans (PP.H1), in East Asians (PP.H2), or with plasma lipids in both populations (PP.H4=10^-6^) at the CETP locus. A posterior probability for a shared genetic signal larger than 0.80 was considered as evidence of colocalization^26^.

To visualise the CETP association with plasma lipids across ancestries, we generated regional association plots of *CETP* using the lipids summary statistics for East Asian and European populations from GLGC. The plots were created with the skyline genomic plotting library (https://gitlab.com/cfinan/skyline) implemented in Python, based on the East Asian and European LD references from UKB.

### Mendelian randomization analysis

To proxy the effect of CETP inhibition we capitalised on *CETP* variants strongly associated with HDL-C in both populations and performed a biomarker weighted drug target MR, by exploring the causal effects of CETP inhibition scaled towards a standard deviation (SD) increase in HDL-C. Despite weighting by an intermediate biomarker, the inference of such a “biomarker” drug target MR analysis is on the protein, not on the potential causality of the intermediate biomarker (**Supplementary Methods** and **Supplemental Figure 1**).

To identify instruments for CETP inhibition, weighted by HDL-C, genetic variants within ±50 kb of the CETP gene (Chr 16:56,995,762-57,017,757, GRCh37) were selected in both populations, based on an F-statistic of at least 15, MAF ≥0.01, and LD-clumped to an r-squared < 0.3 against their respective reference populations. Ancestry specific LD reference matrices were generated by selecting a random subset of 5,000 unrelated Europeans, and the entire subset of East Asians (n=2,000) from UKB. The self-defined East Asian individuals were assigned to the East Asian ancestry group based on principle component analysis, implemented with PC-AiR for the detection of population structure, followed by PC-Relate to account for cryptic relatedness^28^, as described by Giannakopoulou *et al*^29^.

Residual LD was modelled through generalised least squares^30^ implementations of the inverse variance weighted (IVW) and MR-Egger estimators, where the MR-Egger estimator is more robust to the presence of potential horizontal pleiotropy^31^. To further minimise the potential influence of horizontal pleiotropy, we excluded variants with large leverage or outlier statistics and used the Q-statistic to identify possible remaining violations^32^. A model selection framework was applied to select the most appropriate estimator between IVW or MR-Egger for each specific exposure-outcome relationship^32, 33^. This model selection framework, originally developed by Gerta Rücker^34^, utilises the difference in heterogeneity between the IVW Q-statistic and the Egger Q-statistic, preferring the latter model when the difference is larger than 3.84 (i.e., the 97.5% quantile of a Chi-square distribution with 1 degree of freedom). The results were reported as odds ratios (OR) or mean differences (MD) with 95% confidence intervals.

### Interaction test

Potential differences between European and East Asian participants in the drug target MR effects of on-target CETP inhibition were formally tested using interaction tests^35^. Briefly, an interaction effect represents the difference between the ancestry specific MR effects, where the standard error of this difference is equal to the square root of the sum of the variance of the ancestry specific effect estimates. For binary outcomes, where the ancestry specific effect represents an OR, instead of a difference, the interaction effect was calculated as the ratio between the European and East Asian ancestry specific OR (i.e., representing a difference on the logarithmic scale). We additionally applied the interaction test to assess the difference in CETP effects between the East Asian population in our MR study and a previous MR analysis conducted by Millwood *et al.* in China Kadoorie Biobank.

### Multiple testing

The focus of the presented analysis was evaluation of potential differential effects of CETP inhibition between participants of East Asian and European descent. To guard against multiplicity, interaction tests were evaluated against a corrected alpha of 0.05/26 = 1.9×10^-3^, accounting for the 26 evaluated traits. Similarly, comparing the previous analysis by Millwood *et. al.* we had 14 common traits, resulting in a multiplicity corrected interaction p-value of 3.6×10^-3^. We did not apply a similar multiple testing corrected alpha for the ancestry specific findings, and instead focussed on outcomes significant in both ancestries. Focussing on replicated associations resulted in an alpha of 0.050^2^ = 0.0025, and an expected number of false positive results close to zero: 26×0.50^2^=0.065.

### Inferential consideration in biomarker weighted drug target MR

As detailed in Schmidt *et. al.* 2020^36^, Schmidt *et. al*. 2022^37^, and described in the supplementary methods (**Supplementary Methods)**, the inference in biomarker weighted drug target MR is on the drug target itself, not on the downstream biomarker (e.g., HDL-C). Furthermore, the biomarker does not need to cause disease if the drug target affects disease through alternative pathways (i.e., post-translation horizontal pleiotropy) (**Supplemental Figure 1**). We further expand these derivations to show that the biomarker weighted drug target MR will approximate an interaction test of the difference in protein effects, only when the protein effect on the biomarker is equal in both populations (**Supplementary Methods)**. Alternatively, assuming directional concordance of the protein effect on the biomarker, more robust inference will be obtained by applying interaction testing to identify directionally discordant outcome effects.

## Results

### Lack of LDL-C cross-ancestry colocalization at the *CETP* locus

We conducted colocalization analysis to explore potential cross-ancestry colocalization of *CETP* variants associated with LDL-C and HDL-C **(Figure 1, Supplemental Figure 2)**. Comparing the *CETP* HDL-C signal between European and East Asian participants, we observed a high posterior probability (1.00) for a colocalized signal driven by rs183130 (16:g.56991363C>T, GRCh37), which is a known fine-mapped CETP variant in Europeans. Unlike in Europeans, the East-Asian GWAS for LDL-C did not reach genome-wide significance (p-value=6.6×10^-4^) within the *CETP* locus, resulting in a low posterior probability for cross-ancestry colocalization (0.22) **(Figure 1)**.

**Central illustration. Drug target Mendelian randomization framework.** Ancestry specific HDL-C weighted drug target Mendelian randomization (MR) performed in the European (n=1,320,016) and East Asian (n=146,492) populations to assess the effect of CETP inhibition on cardiometabolic biomarkers, cardiovascular and non-cardiovascular traits by leveraging large sample size genome wide association studies (GWAS). To identify instruments in *cis* for CETP inhibition, weighted by HDL-C, genetic variants within ±50 kb of the *CETP* gene were selected in both populations. TG = triglycerides, Apo-A1 = apolipoprotein A, Apo-B = apolipoprotein B, CHD = coronary artery disease, CKD = chronic kidney disease, COPD = chronic obstructive pulmonary disease, CRP = C-reactive protein, CVD = cardiovascular disease, DBP = diastolic blood pressure, HDL-C = high-density lipoprotein cholesterol, HF = heart failure, LDL-C = low-density lipoprotein cholesterol, Lp[a] = lipoprotein a, PAD = peripheral artery disease, SBP = systolic blood pressure, T2D = type 2 diabetes, IS = ischemic stroke, VA = ventricular arrythmia, IH = intracerebral haemorrhage, SH = subarachnoid haemorrhage.

**Figure 1.**
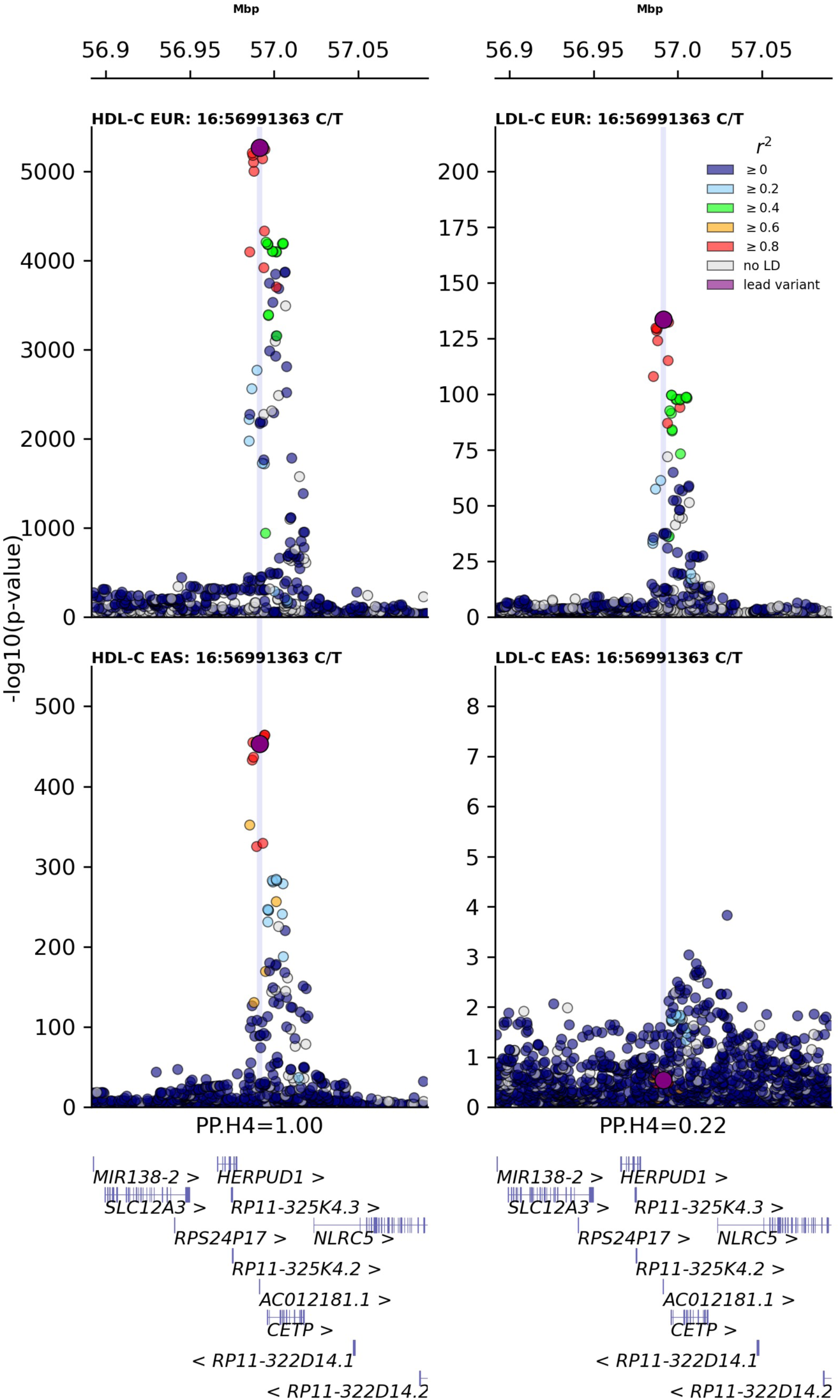
Regional association plots for the *CETP* locus across LDL-C and HDL-C in European (top) and East Asian (bottom) populations. The data were sourced from the global lipids genetics consortium (GLGC), with ancestry specific linkage disequilibrium data obtained from the UK biobank. The y-axis shows the -log_10_(p-values) of the association between each SNP and lipid outcomes. The x-axis shows the chromosomal position (GRCh37). The purple circle shows the European-lead variant rs183130 (16:g.56991363C>T, GRCh37) at the *CETP* locus identified in the GLGC meta-analysis. The colour coding indicates the linkage disequilibrium with the lead fine-mapped European variant based on the UK Biobank European and East Asian reference population. The blue line shows the alignment of the *CETP* signals between lipids, as an indicator of colocalization, reported as posterior probability of both lipids sharing the same causal variant at the *CETP* locus (PP.H4).

### On-target effects of CETP inhibition proxied by HDL-C on biomarkers

Given the lack of LDL-C signal in East Asian participants, we performed a HDL-C weighted drug target MR, scaling *CETP* variants by an SD increase in HDL-C **(Supplemental Table1)**. In European ancestries, lower CETP levels proxied through elevated HDL-C were associated with higher concentrations of Apo-A1, and lower LDL-C, Apo-B, TG, Lp[a], blood pressure, PP, glucose and CRP **(Figure 2)**. In East Asians, we were able to confirm the association between lower CETP and Apo-A1 0.67 g/l (95%CI 0.54; 0.80), TG -0.12 mmol/l (95%CI -0.14; -0.09), Lp[a] -0.25 nmol/l (95%CI -0.44; - 0.07), DBP -0.03 mmHg (95%CI -0.05; -0.01), and glucose -0.04 mmol/l (95%CI -0.07; -0.02).

**Figure 2.**
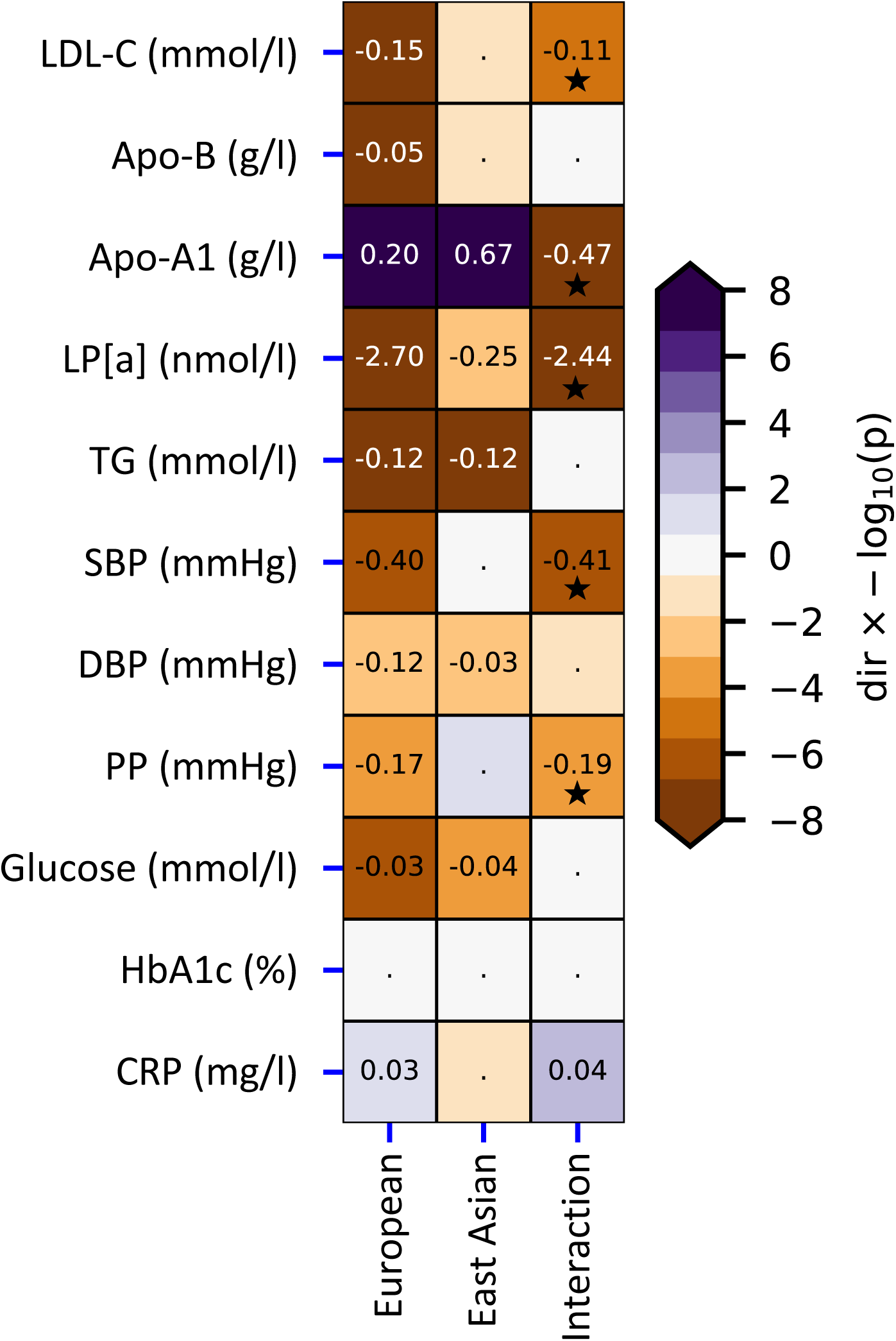
Mendelian randomization effect estimates of lower CETP weighted by HDL-C on biomarkers in East Asian and European populations. The rows represent the plasma biomarker outcomes, with ancestry specific effects presented in the first two columns, and their interaction effect presented in the final column. Cells are annotated by the point estimate (in the indicated units) when these were significant at an alpha of 0.05, or by a point otherwise. Multiplicity correct interaction effects (p-value < 1.9×10^-3^) were additionally annotated by a star symbol. Cells are coloured by the direction of effect (dir) times the -log_10_(p-value), which was truncated to +-8 for display purposes. Apo-A1: apolipoprotein A, Apo-B: apolipoprotein B, Lp[a] lipoprotein a, SBP: systolic blood pressure, DBP: diastolic blood pressure, PP: pulse pressure, CRP: C-reactive protein.

Accounting for multiplicity, we observed significant differences in effect magnitude between ancestries for LDL-C, Apo-A1, Lp[a], SBP, and PP, but not in effect direction **(Figure 2, Supplemental Table 2)**. The LDL-C association in East Asians was -0.04 mmol/L (95%CI -0.09; 0.00, p-value=0.06) compared to -0.15 mmol/L (95%CI -0.16; -0.14, p-value < 1×10^-100^) in Europeans (interaction p-value=6.89×10^-6^). While the effect of CETP inhibition on Apo-A1 was larger in East Asians 0.67 g/l (95%CI 0.54; 0.80) compared to Europeans 0.20 g/l (95%CI 0.20; 0.21) (interaction p-value = 1.01×10^-12^), the CETP effect on Lp[a] was larger in Europeans -2.70 nmol/l (95%CI -3.27; -2.13) compared to East Asians -0.25 nmol/l (95%CI -0.44; -0.07) (interaction p-value = 1.22×10^-15^) (**Supplemental Table 2)**.

### On-target effects of CETP inhibition proxied by HDL-C on clinical outcomes

Lower CETP levels, proxied by HDL-C, was associated with a decreased risk of CHD, angina, HF, intracerebral and subarachnoid haemorrhage in Europeans (**Figure 3**, **Supplemental Table1**). The majority of these associations were replicated in East Asians for: CHD (OR 0.89, 95%CI 0.84; 0.94), angina (OR 0.91, 95%CI 0.84; 0.99), HF (OR 0.85, 95%CI 0.78;0.94), and for intracerebral haemorrhage (OR 0.69, 95%CI 0.55; 0.87). We did not observe significant differences in CETP effect magnitude or direction between the populations (**Figure 3, Supplemental Table 2**).

**Figure 3.**
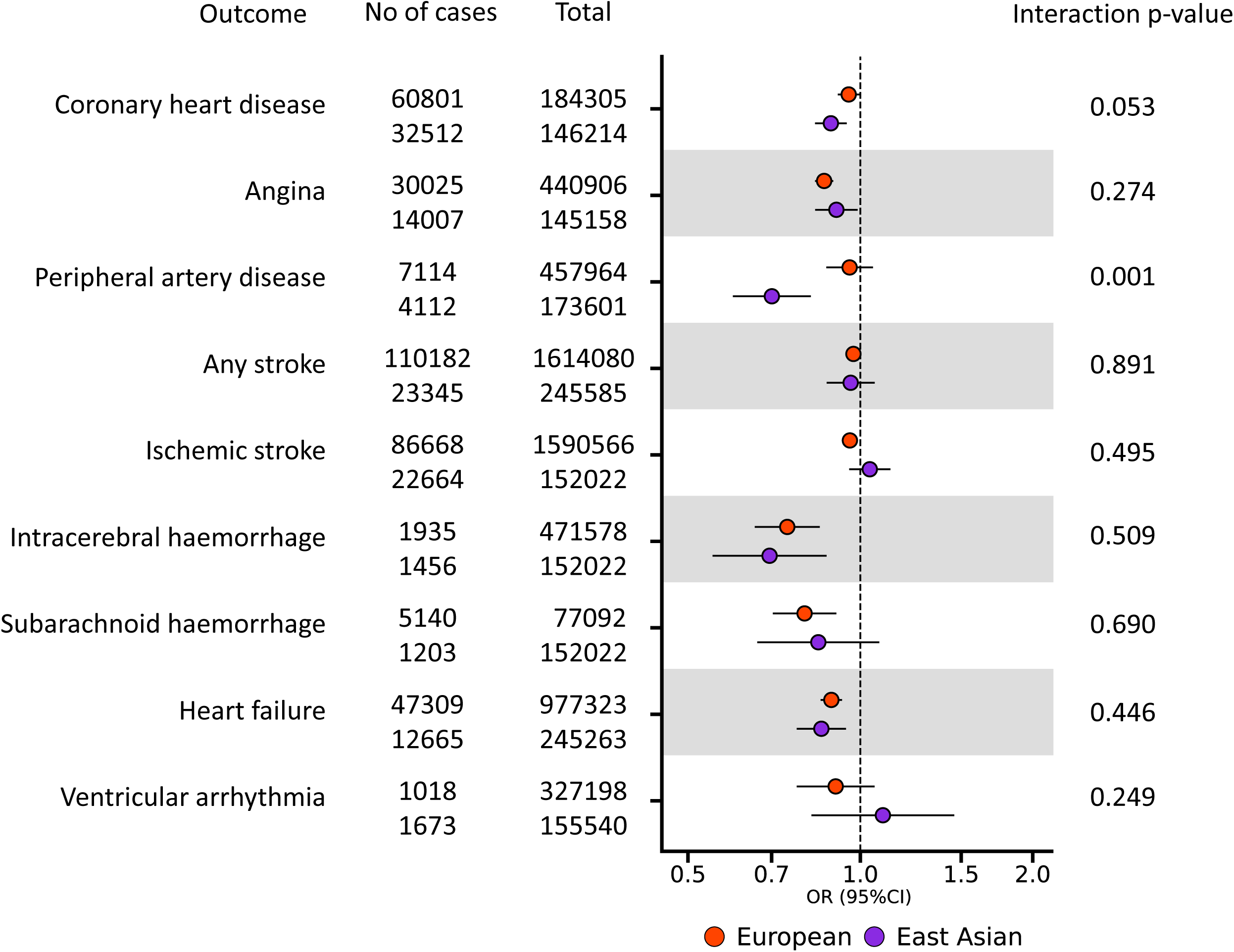
Mendelian randomization effect estimates of lower *CETP* weighted by HDL-C on cardiovascular outcomes in European and East Asian populations. Effect estimates are presented as odds ratios (OR) with 95% confidence intervals (CI) per standard deviation increase in HDL-C. The number of cases and total sample size per outcome is shown on the left. The significance of the interaction between the ancestry specific MR estimates is shown on the right. The multiplicity corrected interaction test alpha was 1.9×10^-3^.

Exploring potential associations with non-cardiovascular outcomes, we observed a protective effect of lower CETP against pneumonia in both Europeans (OR 0.87, 95%CI 0.84; 0.90) and East Asians (OR 0.89, 95%CI 0.81; 0.99) (**Figure 4, Supplemental Table 1**). After accounting for multiplicity, we observed a differential effect of lower CETP on asthma and CKD (interaction p-value < 1.9×10^-3^), with a protective effect in Europeans (asthma: OR 0.95, 95%CI 0.91; 0.99; CKD: OR 0.93, 95%CI 0.90; 0.97), and a risk increasing effect in East Asians (asthma: OR 1.26, 95%CI 1.16; 1.36; CKD: OR 1.31, 95%CI; 1.05; 1.63) (**Figure 4, Supplemental Table 2**).

**Figure 4.**
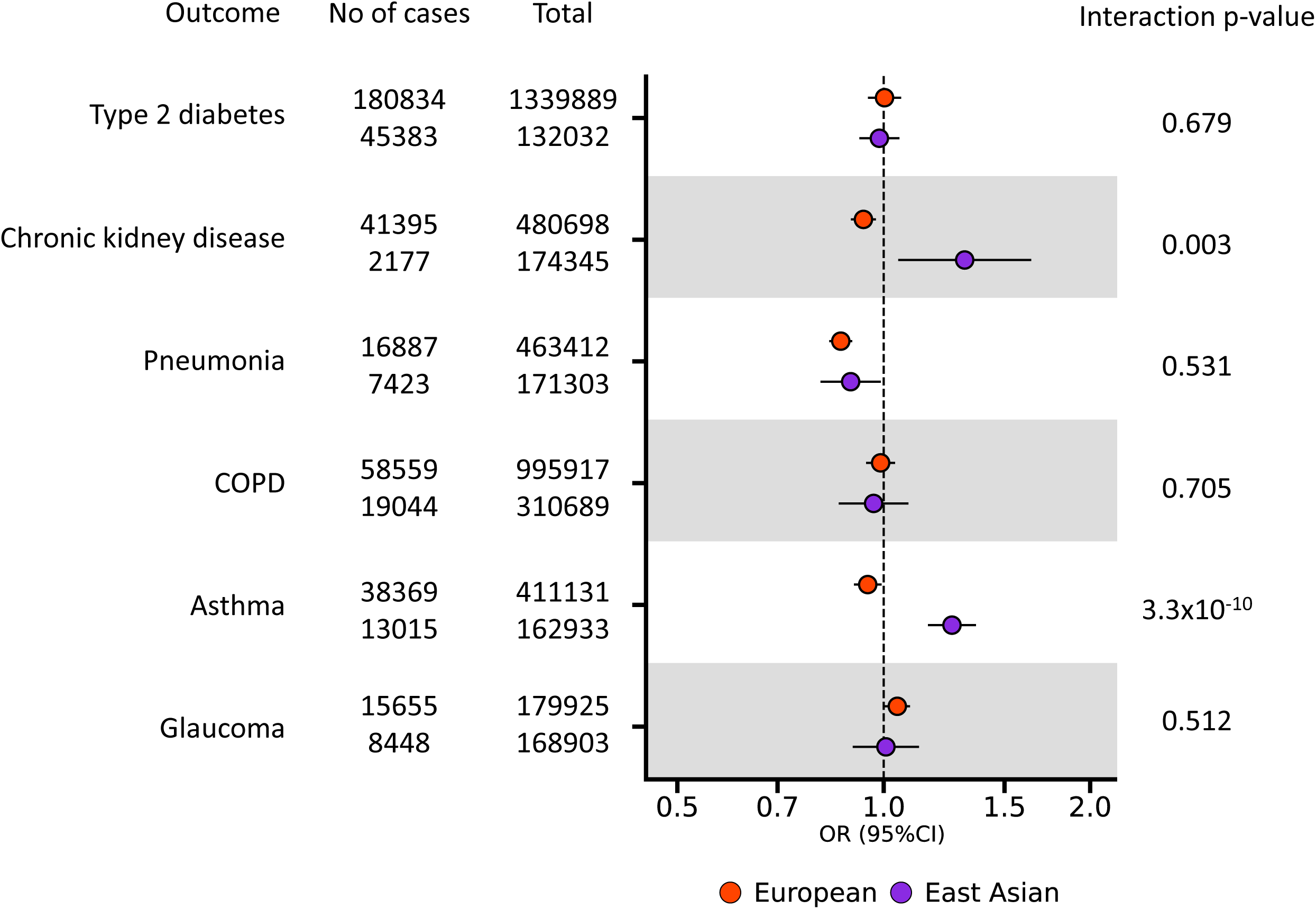
Mendelian randomization effect estimates of lower *CETP* weighted by HDL-C on non-cardiovascular outcomes in European and East Asian ancestries. Effect estimates are presented as odds ratios (OR) with 95% confidence intervals (95%CI) per standard deviation increase in HDL-C (left). The number of cases and total sample size per outcome is shown on the left. The significance of the interaction between the ancestry specific MR estimates is shown on the right. The multiplicity corrected interaction test alpha was 1.9×10^-3^.

### Comparison of CETP effects against Millwood *et al*

We compared the CETP effect, weighted by HDL-C, on cardiometabolic biomarkers and clinical outcomes presented in our current analysis against the results by Millwood *et al.* conducted in China Kadoorie Biobank (**Supplemental Table 3-4**). Despite the larger effect observed by Millwood *et al.* for Apo-A1 1.29 g/l (95%CI 1.04; 1.35) and SBP -0.73 mmHg (95%CI -0.24; -1.21), we did not observe a significant difference of lower CETP levels on these traits. Notably, a difference in effect magnitude was found between the effect of CETP on CHD between the current studies (OR 0.89, 95%CI 0.84;0.94) and the Millwood *et al.* analysis (OR 1.08, 95%CI 0.95;1.22) (interaction p-value= 4.90×10^-3^).

## Discussion

In this study we compared the on-target effect profile of lower CETP levels between individuals of East Asian or European ancestries. Contrary to European populations, where genetic variants in and around *CETP* associate with both HDL-C and LDL-C concentration, in East Asian populations *CETP* variants seem to exclusively affect HDL-C concentration. Using drug-target MR we estimated the on-target effects of lower CETP levels in both populations, which indicated that lower CETP had a larger effect on Apo-A1, LDL-C, LP[a], SBP, and PP in people of European ancestry. Nevertheless, we observed a similar risk decreasing effect of lower CETP levels on CVD outcomes, including CHD, in individuals of East Asian (OR 0.89, 95%CI 0.84; 0.94) and European ancestries (OR 0.95, 95%CI 0.92; 0.99).

Our results are consistent with the previously reported reduction in cardiovascular disease risk observed in the REVEAL clinical trial of anacetrapib^2^, which included a substantial number of participants from China (n=4,314). The effect of anacetrapib on major coronary events in Chinese participants of the REVEAL trial (rate ratio 0.84, 95%CI 0.75; 0.95) was comparable to the aforementioned MR CETP effect on CHD in East Asian: OR 0.89 (95%CI, 0.84; 0.94). This suggests that the absence of a CHD effect in the Millwood *et al.* drug target MR analysis is likely due to the smaller number of participants available to Millwood *et al.* (17,854 had lipid measurement, compared to 146,492 East Asian subjects in our analysis), which can lead to weak-instrument bias towards a neutral effect^38^.

In line with the lack of genetic associations of CETP with LDL-C in individuals of East Asian ancestry, we did not observe a significant effect of lower CETP concentration on LDL-C levels: -0.04 mmol/L (95%CI -0.09; 0.00, p-value=0.06). Randomized controlled trials of anacetrapib conducted in Japanese individuals did however show a decreasing effect of CETP inhibition on LDL-C concentration: −38.0% (95%CI −42.4; −33.7) change from baseline^39^, suggesting that the lack of LDL-C signal observed in our study, as well as that of Millwood et al, is likely limited to the genetic effects of CETP variants on LDL-

C, and does not reflect a fundamental difference in biology of CETP between the ancestries. In line with the results from CETP inhibitor trials, we did not observe differences in the MR effects of lower CETP levels on cardiovascular events between ancestries. Instead, we observed that lower CETP levels decreased the risk of CHD, angina, intracerebral haemorrhage, and heart failure in both ancestry groups. Furthermore, while we did observe some evidence for difference in the effect magnitude of lower CETP levels on biomarkers such as SBP, Apo-A1, and Lp[a], this did not result in directionally opposing effects, at most suggesting a differential amount of CETP inhibition might be considered in East Asian populations.

Considering all the 26 evaluated traits, we only observed a directionally discordant effect for asthma and CKD, where lower CETP levels increased the risk of both diseases in East Asian ancestries, while decreasing the risk in European ancestries. We observed similar risk decreasing effects on asthma and CKD in European centric MR analysis of lower CETP protein concentration^5^. Lower plasma concentrations of CETP have been previously linked to asthma in non-randomized observational studies^40^. A clear biological rationale for CETP involvement in asthma or CKD is however currently absent, prohibiting a straightforward evaluation of the validity of the observed risk increasing effects in individuals of East Asian ancestry. Although approximately 10-30% of participants enrolled in CETP-inhibitor trials are of East Asian ancestry^4, 39, 41^, these studies have not been designed to detect potential differences between ancestry groups, and hence the lack of observed association does not fully rule out a possible ancestry specific risk increasing effects.

The observed comparability in effects of lower CETP levels on CVD outcomes in both European and East Asian populations, despite attenuated effects on CVD risk factors in the later group, argues for the important of a wholistic evaluation of the available evidence in terms of outcomes and traits, as well as in populations, and types of studies. Due to the extensive evidence from CETP inhibitor trials, which specifically recruited participants from East Asian countries, there was strong prior evidence to suggest that CETP should have an effect on CVD in East Asian populations. Without similar supporting evidence from trials, a comparison across different ancestries as presented here – focussing on difference in effect direction instead of effect magnitude, would come to a similar conclusion on the absence of meaningful differences between both populations. The growing genetic data from non-European populations may provide further opportunities to conduct similar analyses, potentially uncovering drug targets overlooked by European centric studies. Above all, the presented results provide yet another reminder to not mistake lack of statistical significance for proof of the null-hypothesis^42^.

## Study limitations

The following potential limitations deserve consideration. First of all, genetic variants from GWAS have a small, presumed cumulative effect size over a lifetime, while pharmacological inhibition of CETP have larger effects, usually prescribed later in life. Consequently, drug target MR estimates may indicate a lifelong effect of perturbing a drug target, which may not be representative of pharmacological interventions at a specific time point and for a shorter period^43^. Although drug target MR does not directly reflect the effect magnitude of the pharmacological intervention, it is a robust indicator of the direction of causal effects^37^. Secondly, we used a biomarker weighted drug target MR approach that does not indicate the possible mediating pathways of the drug target on the disease, but rather reflects the on-target effects drug target perturbation irrespective of the downstream pathway. Thirdly, we note that some residual heterogeneity may reflect design artefacts rather than actual biology. Heterogeneity across biobanks and study cohorts, might occur due to different disease outcome definitions, or participants recruitment criteria, and highlight the need to standardise and prioritise data collection in multi-ancestry cohorts^24^. For example, while East Asian data was predominantly sourced from the BBJ, the European ancestry data represents an amalgamation of distinct European ancestry groups which combined may induce heterogeneity^44^. Rather than design artefacts, observed heterogeneity may reflect influence of distinct environmental settings modifying CETP expression rather than genetic ancestry. Given the various possible sources of heterogeneity, it is important to highlight that we only observed limited differences between the effects profiles of lower CETP levels in European and East Asian ancestries, and instead observed a general tendency for shared beneficial effects of lower CETP levels. We do note however, that due to the relatively limited number of outcomes available in East Asian populations, we were unable to evaluate all relevant clinical outcomes. For example, lower levels of CETP have been linked to increased risk of age-related macular degeneration outcome currently unavailable in genetic studies of East Asian subjects^7^ . Finally, we note that due to the relatively limited amount of data available from East Asian ancestries, there was partial overlap between the exposure data sourced from GLGLC and the outcome GWAS data predominantly sourced from BBJ. As shown by Burgess *et al.*^45^ such partial overlap might cause a limited amount of bias in weak instrument settings, often defined as an F-statistic below 10. It is therefore important to emphasize the instruments were sources from a large number of subjects, based on a minimal F-statistic of 15, where above all the comparability between MR effects in European ancestries and CETP inhibitor trials suggests the impact of any potential weak-instrument bias was minimal.

## Conclusion

In conclusion, lower CETP levels, proxied by HDL-C, had a consistent protective effect against coronary heart disease, angina, heart failure and intracerebral haemorrhage across both ancestries. Therefore, sufficiently potent on-target inhibition of CETP is anticipated to prevent cardiovascular disease in both European and East Asian populations.

## Perspectives

Competencies in medical knowledge: Our findings suggest that CETP inhibition is expected to reduce the risk of cardiovascular disease in individuals of European, as well as East Asian ancestries.

Translational outlook: The use of CETP inhibitors as a potential treatment for coronary heart disease, angina, heart failure and intracerebral haemorrhage can be generalized to the European and East Asian populations.

## Footnotes

The analysis was conducted using Python v.3.7 (GNU Linux). All data needed to evaluate the conclusions in the paper are present in the paper and the Supplement. The software code to create the illustrations and tables have been deposited on UCL figshare.

### Abbreviations

Apo-A1: apolipoprotein
A Apo-B: apolipoprotein
B BBJ: Biobank Japan
CETP: cholesteryl ester transfer protein
CHD: coronary artery disease
CKD: chronic kidney disease
COPD: chronic obstructive pulmonary disease
CRP: C-reactive protein
CVD: cardiovascular disease
DBP: diastolic blood pressure
GBMI: Global Biobank Meta-analysis Initiative
GLGC: Global Lipids Genetics Consortium
GWAS: genome-wide association study
HDL-C: high-density lipoprotein cholesterol
HF: heart failure
LDL-C: low-density lipoprotein cholesterol
Lp[a]: lipoprotein a
PAD: peripheral artery disease
SBP: systolic blood pressure
T2D: type 2 diabetes
UKB: UK biobank

## Data Availability

All data produced in the present work are contained in the manuscript

## Acknowledgements

We thank the Medical Research Council Doctoral Training Programme for funding this research. We additionally thank the participants of the Global lipids Genetics Consortium, UK Biobank, and Biobank Japan, Global Biobank Meta-analysis Initiative.

## Notes

### Competing Interest Statement

AFS and CF have received unrestricted funding from NewAmsterdam, which is currently developing the CETP-inhibitor obicetrapib. The views expressed in this study are the personal views of MGM and do not represent the views of her current employer, the European Medicines Agency. All the authors declare no other competing interests.

### Funding Statement

This research was funded by the Medical Research Council Doctoral Training Programme, grant MR/NO13867/1, awarded to DD. KK is supported by the European Research Council under the European Union Horizon 2020 research and innovation program grant 948561. ADH is a NIHR Senior Investigator and supported by the UKRI-NIHR grant MR/V033867/1 for the Multimorbidity Mechanism and Therapeutics Research Collaboration. CF acknowledges support from UCL BHF Research Accelerator grant AA/18/34223, and MR/V033867/1. AFS is supported by the BHF grants PG/18/5033837, PG/22/10989, AA/18/34223, and MR/V033867/1.

### Author Declarations

The study used (or will use) ONLY openly available human data that were originally located at: https://pan.ukbb.broadinstitute.org/, http://www.nealelab.is/uk-biobank, https://www.nature.com/articles/s41588-021-00931-x, https://www.sciencedirect.com/science/article/pii/S0002929718303203?via%3Dihub, https://www.globalbiobankmeta.org/, https://www.nature.com/articles/s41588-020-0640-3#article-info, https://www.nature.com/articles/ng.3396, https://www.ncbi.nlm.nih.gov/pubmed/31959993, https://www.nature.com/articles/s41588-022-01058-3, https://www.nature.com/articles/s41588-021-00852-9#Abs1, https://www.nature.com/articles/s41467-019-13690-5

## References

1. Barter PJ, Hopkins GJ, Calvert GD. Transfers and exchanges of esterified cholesterol between plasma lipoproteins. Biochemical Journal. 1982;208(1):1–7. doi:10.1042/bj2080001

2. HPS3/TIMI55–REVEAL Collaborative Group, Bowman L, Hopewell JC, et al. Effects of Anacetrapib in Patients with Atherosclerotic Vascular Disease. N Engl J Med. 2017;377(13):1217–1227.

3. Barter PJ, Caulfield M, Eriksson M, et al. Effects of torcetrapib in patients at high risk for coronary events. N Engl J Med. 2007;357(21):2109–2122.

4. Lincoff AM, Michael Lincoff A, Nicholls SJ, et al. Evacetrapib and Cardiovascular Outcomes in High-Risk Vascular Disease. New England Journal of Medicine. 2017;376(20):1933–1942. doi:10.1056/nejmoa1609581

5. Schmidt AF, Hunt NB, Gordillo-Marañón M, et al. Cholesteryl ester transfer protein (CETP) as a drug target for cardiovascular disease. Nat Commun. 2021;12(1):5640.

6. Sofat R, Hingorani AD, Smeeth L, et al. Separating the mechanism-based and off-target actions of cholesteryl ester transfer protein inhibitors with CETP gene polymorphisms. Circulation. 2010;121(1):52–62.

7. Cupido AJ, Reeskamp LF, Hingorani AD, et al. Joint Genetic Inhibition of PCSK9 and CETP and the Association With Coronary Artery Disease: A Factorial Mendelian Randomization Study. JAMA Cardiol. 2022;7(9):955–964.

8. Millwood IY, Bennett DA, Holmes MV, et al. Association of CETP Gene Variants With Risk for Vascular and Nonvascular Diseases Among Chinese Adults. JAMA Cardiol. 2018;3(1):34–43.

9. Zhong S, Sharp DS, Grove JS, et al. Increased coronary heart disease in Japanese-American men with mutation in the cholesteryl ester transfer protein gene despite increased HDL levels. J Clin Invest. 1996;97(12):2917–2923.

10. Curb JD, Abbott RD, Rodriguez BL, et al. A prospective study of HDL-C and cholesteryl ester transfer protein gene mutations and the risk of coronary heart disease in the elderly. J Lipid Res. 2004;45(5):948–953.

11. Barter PJ, Brewer HB Jr, Chapman MJ, Hennekens CH, Rader DJ, Tall AR. Cholesteryl ester transfer protein: a novel target for raising HDL and inhibiting atherosclerosis. Arterioscler Thromb Vasc Biol. 2003;23(2):160–167.

12. Sakaue S, Kanai M, Tanigawa Y, et al. A cross-population atlas of genetic associations for 220 human phenotypes. Nat Genet. 2021;53(10):1415–1424.

13. Graham SE, Clarke SL, Wu KHH, et al. The power of genetic diversity in genome-wide association studies of lipids. Nature. 2021;600(7890):675–679.

14. Wain LV, Vaez A, Jansen R, et al. Novel Blood Pressure Locus and Gene Discovery Using Genome-Wide Association Study and Expression Data Sets From Blood and the Kidney. Hypertension. Published online July 24, 2017. doi:10.1161/HYPERTENSIONAHA.117.09438

15. Chen J, Spracklen CN, Marenne G, et al. The trans-ancestral genomic architecture of glycemic traits. Nat Genet. 2021;53(6):840–860.

16. Ligthart S, Vaez A, Võsa U, et al. Genome Analyses of >200,000 Individuals Identify 58 Loci for Chronic Inflammation and Highlight Pathways that Link Inflammation and Complex Disorders. Am J Hum Genet. 2018;103(5):691–706.

17. Nikpay M, Goel A, Won HH, et al. A comprehensive 1,000 Genomes-based genome-wide association meta-analysis of coronary artery disease. Nat Genet. 2015;47(10):1121–1130.

18. Mishra A, Malik R, Hachiya T, et al. Stroke genetics informs drug discovery and risk prediction across ancestries. Nature. 2022;611(7934):115–123.

19. Shah S, Henry A, Roselli C, et al. Genome-wide association and Mendelian randomisation analysis provide insights into the pathogenesis of heart failure. Nat Commun. 2020;11(1):163.

20. Mahajan A, Spracklen CN, Zhang W, et al. Multi-ancestry genetic study of type 2 diabetes highlights the power of diverse populations for discovery and translation. Nat Genet. 2022;54(5):560–572.

21. Wuttke M, Li Y, Li M, et al. A catalog of genetic loci associated with kidney function from analyses of a million individuals. Nat Genet. 2019;51(6):957–972.

22. Craig JE, Han X, Qassim A, et al. Multitrait analysis of glaucoma identifies new risk loci and enables polygenic prediction of disease susceptibility and progression. Nat Genet. 2020;52(2):160–166.

23. Bakker MK, van der Spek RAA, van Rheenen W, et al. Genome-wide association study of intracranial aneurysms identifies 17 risk loci and genetic overlap with clinical risk factors. Nat Genet. 2020;52(12):1303–1313.

24. Zhou W, Kanai M, Wu KHH, et al. Global Biobank Meta-analysis Initiative: Powering genetic discovery across human disease. Cell Genomics. 2022;2(10). doi:10.1016/j.xgen.2022.100192

25. Website. https://pan.ukbb.broadinstitute.org. 2020.

26. Giambartolomei C, Vukcevic D, Schadt EE, et al. Bayesian test for colocalisation between pairs of genetic association studies using summary statistics. PLoS Genet. 2014;10(5):e1004383.

27. Wallace C. Eliciting priors and relaxing the single causal variant assumption in colocalisation analyses. PLoS Genet. 2020;16(4):e1008720.

28. Conomos MP, Miller MB, Thornton TA. Robust inference of population structure for ancestry prediction and correction of stratification in the presence of relatedness. Genet Epidemiol. 2015;39(4):276–293.

29. Giannakopoulou O, Lin K, Meng X, et al. The Genetic Architecture of Depression in Individuals of East Asian Ancestry: A Genome-Wide Association Study. JAMA Psychiatry. 2021;78(11):1258–1269.

30. Burgess S, Thompson SG. Mendelian Randomization: Methods for Causal Inference Using Genetic Variants. CRC Press; 2021.

31. Burgess S, Butterworth A, Thompson SG. Mendelian randomization analysis with multiple genetic variants using summarized data. Genet Epidemiol. 2013;37(7):658–665.

32. Bowden J, Del Greco M F, Minelli C, Davey Smith G, Sheehan N, Thompson J. A framework for the investigation of pleiotropy in two-sample summary data Mendelian randomization. Stat Med. 2017;36(11):1783–1802.

33. Bowden J, Spiller W, Del Greco M F, et al. Improving the visualization, interpretation and analysis of two-sample summary data Mendelian randomization via the Radial plot and Radial regression. International Journal of Epidemiology. 2018;47(4):1264–1278. doi:10.1093/ije/dyy101

34. Rucker G, Schwarzer G, Carpenter JR, Binder H, Schumacher M. Treatment-effect estimates adjusted for small-study effects via a limit meta-analysis. Biostatistics. 2011;12(1):122–142. doi:10.1093/biostatistics/kxq046

35. Altman DG, Bland JM. Interaction revisited: the difference between two estimates. BMJ. 2003;326(7382):219.

36. Schmidt AF, Finan C, Gordillo-Marañón M, et al. Genetic drug target validation using Mendelian randomisation. Nat Commun. 2020;11(1):1–12.

37. Schmidt AF, Hingorani AD, Finan C. Human Genomics and Drug Development. Cold Spring Harb Perspect Med. 2022;12(2). doi:10.1101/cshperspect.a039230

38. Burgess S, Thompson SG. Avoiding bias from weak instruments in Mendelian randomization studies. Int J Epidemiol. 2011;40(3):755–764.

39. Lipid-modifying efficacy and tolerability of anacetrapib added to ongoing statin therapy in Japanese patients with dyslipidemia. Atherosclerosis. 2017;261:69–77.

40. Xiao J, Lu S, Wang X, et al. Serum Proteomic Analysis Identifies SAA1, FGA, SAP, and CETP as New Biomarkers for Eosinophilic Granulomatosis With Polyangiitis. Front Immunol. 2022;13:866035.

41. Nicholls SJ, Ditmarsch M, Kastelein JJ, et al. Lipid lowering effects of the CETP inhibitor obicetrapib in combination with high-intensity statins: a randomized phase 2 trial. Nat Med. 2022;28(8):1672–1678.

42. Altman DG, Bland JM. Absence of evidence is not evidence of absence. BMJ. 1995;311(7003):485.

43. Gill D, Walker VM, Martin RM, Davies NM, Tzoulaki I. Comparison with randomized controlled trials as a strategy for evaluating instruments in Mendelian randomization. Int J Epidemiol. 2020;49(4):1404–1406.

44. Novembre J, Johnson T, Bryc K, et al. Genes mirror geography within Europe. Nature. 2008;456(7218):98.

45. Burgess S, Davies NM, Thompson SG. Bias due to participant overlap in two-sample Mendelian randomization. Genet Epidemiol. 2016;40(7):597–608.

