## Supplementary methods figures and tables for "Comparing the effect profile of CETP in individuals of East Asian and European ancestries": Supplement copy.docx

### Online Appendix

**Supplementary Methods.** Supplementary methods.

**Supplemental Figure 1**. Drug target Mendelian randomisation pathways.

**Supplemental Figure 2**. Regional association plots for the *CETP* locus across lipid traits (LDL-C, HDL-C, nonHDL-C, TG, TC) in the European and East Asian populations of GLGC.

**Supplemental Table 1.** East Asian and European GWAS outcome datasets used in cis-Mendelian randomisation analysis.

**Supplemental Table 2.** CETP cis-Mendelian randomisation effect estimates on biomarkers and clinical outcomes in the East Asian and European population.

**Supplemental Table 3**. Interaction test of CETP cis-Mendelian randomisation effect estimates on biomarkers and clinical outcomes between the East Asian and European population.

**Supplemental Table 4**. Baseline characteristics of biomarkers across European (UKB), East Asian (this study) and Chinese populations (Millwood *et al.*).

**Supplemental Table 5**. Comparison of CETP effects on clinical outcomes and biomarkers with Millwood *et al.* in the East Asian population.

**Supplementary methods**

**Inference in a biomarker drug target Mendelian randomization analysis**

Following Schmidt *et al.* 2020 and 2022, the graph in **eFigure 1** depicts the data generating model of a drug target Mendelian randomization (MR). Here the absence of an arc between the genetic variants $\boldsymbol{G}$ and the outcome $\boldsymbol{D}$ ensure there is pre-translational horizontal pleiotropy, which would otherwise cause bias of the drug target MR effect of the protein $\boldsymbol{P}$ on the outcome. This protein drug target effect can be referred to as:

$$\omega=\mu\theta+\phi_{\boldsymbol{P}}, eq. 1$$

which consists of the direct effect $\mu\theta$mediated by biomarker $\boldsymbol{X}$, and the indirect effect $\phi_{\boldsymbol{P}}$ a protein might have through a pathway (or pathways) side-stepping $\boldsymbol{X}$. Depending on the application, there might be multiple intermediate biomarkers, resulting in a straightforward expansion of the equation 1.

Because there are confounding factors $\boldsymbol{U,}$ which are a common cause for both $\boldsymbol{P}$ and $\boldsymbol{D}$, simply regressing $\boldsymbol{D}$ on $\boldsymbol{P}$ is not expected to provide an unbiased estimate of $\omega$. Instead, given that the genetic effects on the outcome and the protein are unaffected by confounders, MR can be employed, where the fraction of the genetic effect on the outcome by the genetic effect on the protein results in the intended estimate:

$$\omega=\frac{\tilde{\delta}\left( \mu\theta+\phi_{\boldsymbol{P}} \right)}{\tilde{\delta}},$$

$$=\mu\theta+\phi_{\boldsymbol{P}} .$$

While there is a growing resource on genetic protein associations, sufficient information on $\tilde{\delta}$ might not always be available. Instead, in some cases (e.g., lipids) there might be more information and data on the genetic effect on a biomarker, which is known to be affected by the protein (that is $\mu$). In these cases, a biomarker weighted (bw) drug target MR analysis can be calculated, by replacing $\tilde{\delta}$ with the genetic association on the biomarker:

$$\omega_{bw}=\frac{\tilde{\delta}\left( \mu\theta+\phi_{\boldsymbol{P}} \right)}{\tilde{\delta}\mu},=\frac{1}{\mu}\omega.$$

Clearly, $\omega_{bw}$ is a bias estimand of $\omega$, however assuming sufficient detail is available on the sign of $\mu$, that is information on whether the protein increases or decreases the biomarker concentration, $\omega_{bw}$ can provide key information on the anticipated effect direction of $\omega$. Furthermore, given that $\omega_{bw}=0 \Leftrightarrow\omega=0$ a biomarker weighted drug target MR provides a valid null-hypothesis test of $\omega$ irrespective of the amount of bias due to $\frac{1}{\mu}$.

**Interaction effects using biomarker drug target Mendelian randomization analyses**

Given two distinct populations, say people of European descent and people from East Asian descent, one might be interested in determining to what extent there is a difference in their drug-target effect on the same outcome. In the presence of confounding (eFigure 1) and genetic information on the protein expression in both populations, this can be estimated through a drug target MR:

$$\omega_{j}-\omega_{k}=\left( \mu_{j}\theta_{j}+{\phi_{\boldsymbol{P}}}_{j} \right)-\left( \mu_{k}\theta_{k}+\phi_{\boldsymbol{P}_{k}} \right), eq. 2$$

here $j$ and $k$ represent effects from eFigure 1 for two non-overlapping subgroups, such as European and East Asian participants, respectively. An interaction test for $\omega_{j}-\omega_{k}\neq0$ would provide evidence for a difference.

In the absence of information on protein expression in both populations, one could consider conducting a biomarker weighted drug target MR in both populations and using this to determine the difference in effects between two populations. Given that a biomarker weighted MR provides a biased estimate of $\omega$, one needs to additionally assume that the amount of bias in both populations is equal, we need to assume that $\theta_{j}=\theta_{k}$. To see this, let us assume there is no difference between the protein effect on the outcome in both populations, which is:

$$\omega_{j}-\omega_{k}=0.$$

Furthermore, if we assume (as implicitly above) $\theta_{j},\theta_{k}\neq0$, then the biomarker weighted drug target analysis becomes:

$$\omega_{bw_{j}}-{\omega_{bw}}_{k}=\frac{\theta_{j}\omega_{j}-\theta_{k}\omega_{k}}{\theta_{j}\theta_{k}},$$

Clearly, this can only equal zero when $\theta_{j}=\theta_{k}$.

Biomarker weighted drug target MR can be used to obtain a valid null-hypothesis of $\omega_{j}-\omega_{k}\neq0$, if we assume that the protein effect on the downstream biomarker is equal in both populations. In the absence of an exact agreement between $\theta_{j}$ and $\theta_{k}$, the false positive (i.e., type 1 error) rate of the interaction tests will be inflated proportional to the difference $\theta_{j}-\theta_{k}$. Depending on the application, $\theta_{j}=\theta_{k}$ might be too strong an assumption to make, instead if we are more comfortable assuming the sign of $\theta_{j}$ and $\theta_{k}$is the same (i.e., that a unit increase in the protein does not increase the biomarker in one population, while decreasing in the second), more robust interaction tests can be obtained by focussing on directional discordance between populations. Therefore, focussing on direction of effects might offer a more robust interpretation.

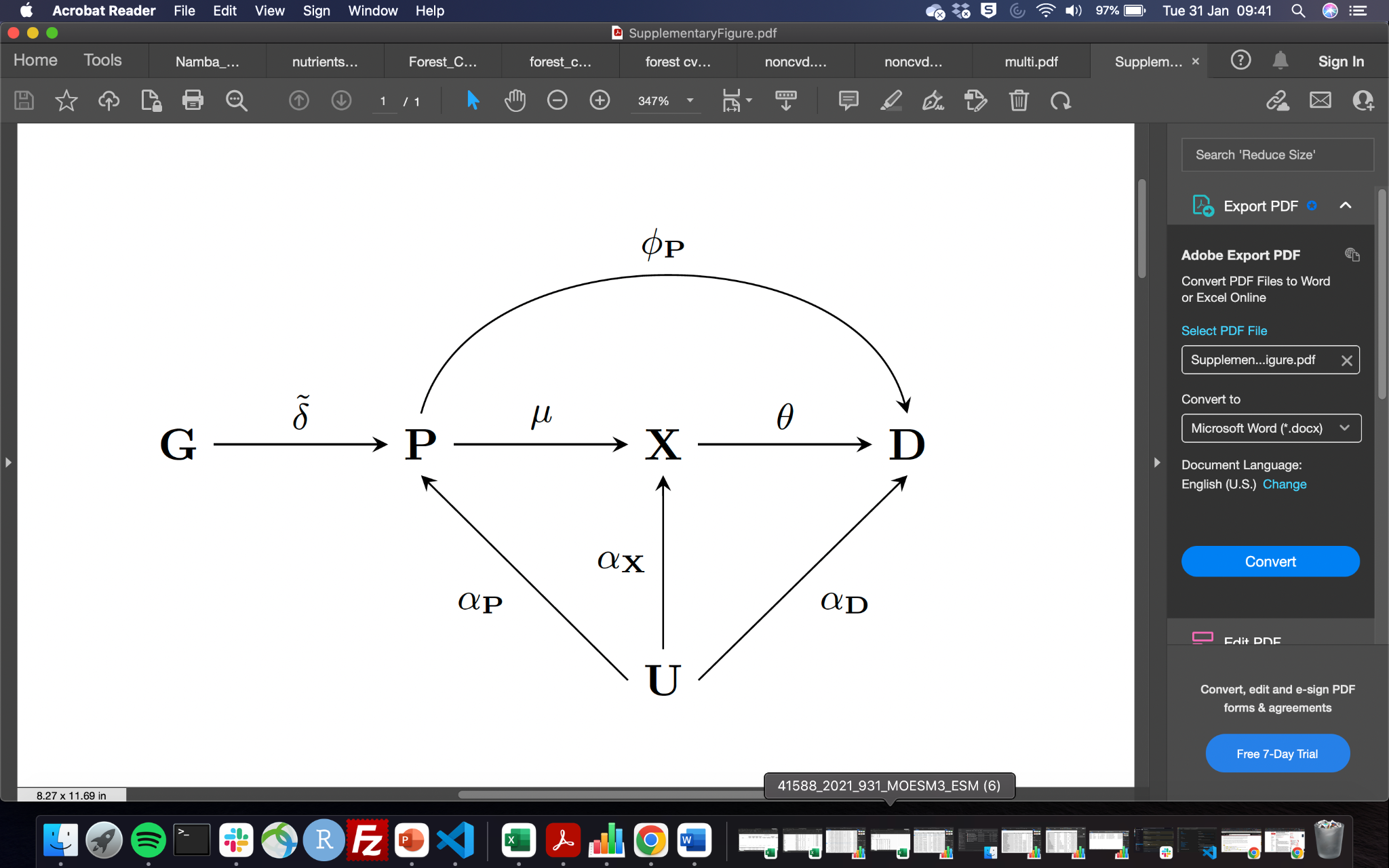

**Supplemental Figure 1. Drug target Mendelian randomisation pathways**. Nodes are presented in bold face, with **G** representing a genetic variant, $\boldsymbol{P}$ a protein drug target, $\boldsymbol{X}$ a biomarker, $\boldsymbol{D}$ the outcome, and $\boldsymbol{U}$ (potentially unmeasured) common causes of both $\boldsymbol{P}$, $\boldsymbol{X}$, $\boldsymbol{D}$. Labelled paths represent the effects between nodes.

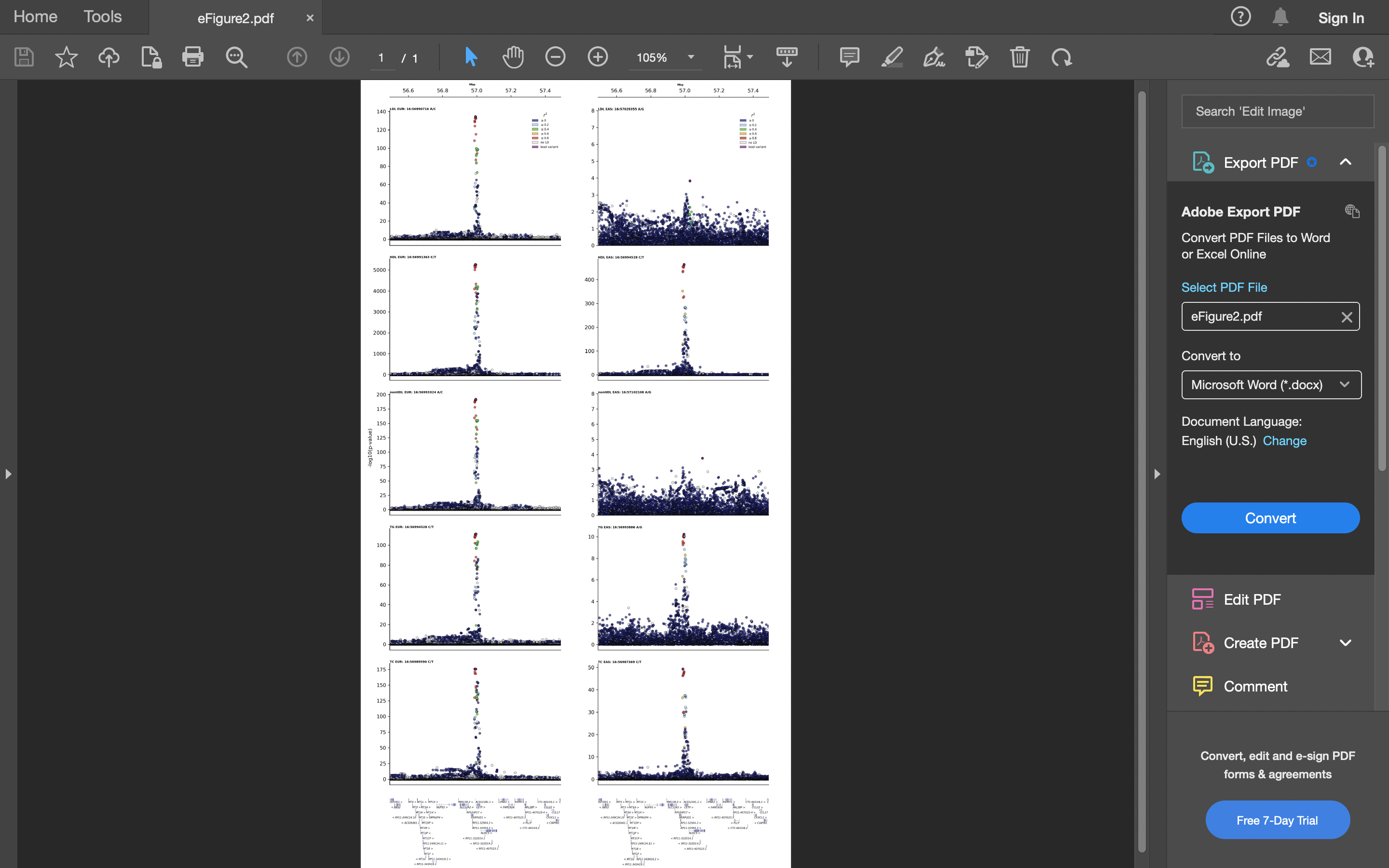

**Supplemental Figure 2. Regional association plots for the *CETP* locus across lipid traits (LDL-C, HDL-C, nonHDL-C, TG, TC) in the European (left) and East Asian (right) populations of GLGC.** The y-axes show the -log10(p-values) of the association between each SNP and lipid outcomes. The x-axes show the chromosomal position (GRCh37). The purple circle shows the lead SNP in each region. The colour coding indicates the linkage disequilibrium with the lead SNP based on the UK Biobank European and East Asian reference population.

**Supplemental Table 1. East Asian and European GWAS outcome datasets used in *cis*-Mendelian randomisation analysis.**

| **Outcomes** | **Ancestry** | **No of cases** | **Total sample size** | **Database** | **URL** |
| --- | --- | --- | --- | --- | --- |
| Apoprotein A1 | East Asian | 0 | 2325 | Pan-UK Biobank | <https://pan.ukbb.broadinstitute.org/> |
|  | European | 0 | 361194 | Pan-UK Biobank | <http://www.nealelab.is/uk-biobank> |
| Apoprotein B | East Asian | 0 | 2553 | Pan-UK Biobank | <https://pan.ukbb.broadinstitute.org/> |
|  | European | 0 | 361194 | Pan-UK Biobank | <http://www.nealelab.is/uk-biobank> |
| Angina | East Asian | 14007 | 145158 | Biobank Japan | <https://www.nature.com/articles/s41588-021-00931-x> |
|  | European | 30025 | 440906 | FinGen + UK Biobank | <https://www.nature.com/articles/s41588-021-00931-x> |
| Asthma | East Asian | 13015 | 162933 | Biobank Japan | <https://www.nature.com/articles/s41588-021-00931-x> |
|  | European | 38369 | 411131 | FinGen + UK Biobank | <https://www.nature.com/articles/s41588-021-00931-x> |
| C-Reactive Protein | East Asian | 0 | 83025 | Biobank Japan | <https://www.nature.com/articles/s41588-021-00931-x> |
|  | European | 0 | 204402 | Ligthart et al., 2018 | <https://www.sciencedirect.com/science/article/pii/S0002929718303203?via%3Dihub> |
| Chronic obstructive pulmonary | East Asian | 19044 | 310689 | GBMI | <https://www.globalbiobankmeta.org/> |
|  | European | 58559 | 995917 | GBMI | <https://www.globalbiobankmeta.org/> |
| Coronary heart disease | East Asian | 32512 | 146214 | Biobank Japan | <https://www.nature.com/articles/s41588-020-0640-3#article-info> |
|  | European | 60801 | 184305 | CARDIoGRAMplusC4D | <https://www.nature.com/articles/ng.3396> |
| Type 2 diabetes | East Asian | 45383 | 132032 | Biobank Japan | <https://www.nature.com/articles/s41588-021-00931-x> |
|  | European | 180834 | 1339889 | Mahjan et al. | <https://www.nature.com/articles/s41588-022-01058-3> |
| Diastolic blood pressure | East Asian | 0 | 145515 | Biobank Japan | <https://www.nature.com/articles/s41588-021-00931-x> |
|  | European | 0 | 757601 | ICBP | <https://www.nature.com/articles/s41588-018-0205-x> |
| Glaucoma | East Asian | 8448 | 168903 | Biobank Japan | <https://www.nature.com/articles/s41588-021-00931-x> |
|  | European | 15655 | 179925 | Craig et al., 2020 | <https://www.ncbi.nlm.nih.gov/pubmed/31959993> |
| Glucose | East Asian | 0 | 133336 | Biobank Japan | <https://www.nature.com/articles/s41588-021-00931-x> |
|  | European | 0 | 196991 | MAGIC | <https://www.nature.com/articles/s41588-021-00852-9#Abs1> |
| HbA1c | East Asian | 0 | 71221 | Biobank Japan | <https://www.nature.com/articles/s41588-021-00931-x> |
|  | European | 0 | 196991 | MAGIC | <https://www.nature.com/articles/s41588-021-00852-9#Abs1> |
| HDL-cholesterol | East Asian | 0 | 74970 | Biobank Japan | <https://www.nature.com/articles/s41588-021-00931-x> |
|  | European | 0 | 115078 | UK Biobank | <https://gwas.mrcieu.ac.uk/datasets/?gwas_id__icontains=met-d> |
| Heart failure | East Asian | 12665 | 245263 | GBMI | <https://www.globalbiobankmeta.org/> |
|  | European | 47309 | 977323 | HERMES | <https://www.nature.com/articles/s41467-019-13690-5> |
| Intracerebral haemorrhage | East Asian | 1456 | 152022 | Biobank Japan | <https://www.nature.com/articles/s41588-021-00931-x> |
|  | European | 1935 | 471578 | FinGen + UK Biobank | <https://www.nature.com/articles/s41588-021-00931-x> |
| Ischemic stroke | East Asian | 22664 | 152022 | Biobank Japan | <https://www.nature.com/articles/s41588-021-00931-x> |
|  | European | 86668 | 1590566 | GIGASTROKE | <https://www.nature.com/articles/s41586-022-05165-3> |
| LDL-cholesterol | East Asian | 0 | 72866 | Biobank Japan | <https://www.nature.com/articles/s41588-021-00931-x> |
|  | European | 0 | 361194 | UK Biobank | <http://www.nealelab.is/uk-biobank> |
| Lipoprotein(a) | East Asian | 0 | 2275 | Pan-UK Biobank | <https://pan.ukbb.broadinstitute.org/> |
|  | European | 0 | 361194 | UK Biobank | <http://www.nealelab.is/uk-biobank> |
| Peripheral arterial disease | East Asian | 4112 | 173601 | Biobank Japan | <https://www.nature.com/articles/s41588-021-00931-x> |
|  | European | 7114 | 457964 | FinGen + UK Biobank | <https://www.nature.com/articles/s41588-021-00931-x> |
| Pneumonia | East Asian | 7423 | 171303 | Biobank Japan | <https://www.nature.com/articles/s41588-021-00931-x> |
|  | European | 16887 | 463412 | FinGen + UK Biobank | <https://www.nature.com/articles/s41588-021-00931-x> |
| Pulse pressure | East Asian | 0 | 145445 | Biobank Japan | <https://www.nature.com/articles/s41588-021-00931-x> |
|  | European | 0 | 757601 | ICBP | <https://www.nature.com/articles/s41588-018-0205-x> |
| Any stroke | East Asian | 23345 | 245585 | GBMI | <https://www.globalbiobankmeta.org/> |
|  | European | 110182 | 1614080 | GIGASTROKE | <https://www.nature.com/articles/s41586-022-05165-3> |
| Subarachnoid haemorrhage | East Asian | 1203 | 152022 | Biobank Japan | <https://www.nature.com/articles/s41588-021-00931-x> |
|  | European | 5140 | 77092 | Bakker et al., 2022 | [www.nature.com/articles/s41588-020-00725-7](http://www.nature.com/articles/s41588-020-00725-7) |
| Systolic blood pressure | East Asian | 0 | 145505 | Biobank Japan | <https://www.nature.com/articles/s41588-021-00931-x> |
|  | European | 0 | 757601 | ICBP | <https://www.nature.com/articles/s41588-018-0205-x> |
| Total cholesterol | East Asian | 0 | 135808 | Biobank Japan | <https://www.nature.com/articles/s41588-021-00931-x> |
|  | European | 0 | 361194 | UK Biobank | <http://www.nealelab.is/uk-biobank> |
| Triglycerides | East Asian | 0 | 111667 | Biobank Japan | <https://www.nature.com/articles/s41588-021-00931-x> |
|  | European | 0 | 115078 | UK Biobank | <http://www.nealelab.is/uk-biobank> |
| Ventricular arrhythmia | East Asian | 1673 | 155540 | Biobank Japan | <https://www.nature.com/articles/s41588-021-00931-x> |
|  | European | 1018 | 327198 | FinGen + UK Biobank | <https://www.nature.com/articles/s41588-021-00931-x> |
| Chronic kidney disease | East Asian | 2117 | 174345 | Biobank Japan | <https://www.nature.com/articles/s41588-021-00931-x> |
|  | European | 41395 | 480698 | Wuttke et al., 2019 | <https://www.nature.com/articles/s41588-019-0407-x> |

**Supplemental Table 2. CETP *cis*-Mendelian randomisation effect estimates on biomarkers and clinical outcomes in the East Asian and European population.**

| **Outcome** | **Ancestry** | **MR estimate (OR/MD)** | **se** | **p-value** | **n variants** | **model** | **Heterogeneity p-value** | **Q-statistic** | **unit** |
| --- | --- | --- | --- | --- | --- | --- | --- | --- | --- |
| Angina | East Asian | 0.91 (0.84; 0.99) | 0.042 | 2.1×10⁻² | 37 | IVW | 0.202 | 42.813 | OR |
|  | European | 0.86 (0.84; 0.89) | 0.017 | 1.0×10⁻¹⁰⁰ | 61 | IVW | 0.246 | 67.115 | OR |
| Any stroke | East Asian | 0.96 (0.88; 1.06) | 0.048 | 4.1×10⁻¹ | 32 | MR Egger | 0.013 | 49.671 | lOR |
|  | European | 0.97 (0.95; 1.00) | 0.013 | 3.9×10⁻² | 35 | IVW | 0.15 | 42.509 | OR |
| Apo-A1 | East Asian | 0.67 (0.54; 0.80) | 0.066 | 1.0×10⁻¹⁰⁰ | 37 | IVW | <0.001 | 73.108 | g/l |
|  | European | 0.20 (0.20; 0.21) | 0.001 | 1.0×10⁻¹⁰⁰ | 48 | MR Egger | <0.001 | 125.445 | g/l |
| Apo-B | East Asian | -0.18 (-0.44; 0.07) | 0.130 | 1.6×10⁻¹ | 36 | MR Egger | 0.054 | 48.213 | g/l |
|  | European | -0.05 (-0.05; -0.05) | 0.001 | 1.0×10⁻¹⁰⁰ | 54 | MR Egger | <0.001 | 105.830 | g/l |
| Asthma | East Asian | 1.26 (1.16; 1.36) | 0.039 | 5.8×10⁻⁹ | 36 | IVW | <0.001 | 82.335 | OR |
|  | European | 0.95 (0.91; 0.99) | 0.022 | 1.4×10⁻² | 58 | MR Egger | 0.136 | 67.712 | OR |
| CHD | East Asian | 0.89 (0.84; 0.94) | 0.031 | 1.2×10⁻⁴ | 32 | IVW | 0.211 | 37.010 | OR |
|  | European | 0.95 (0.92; 0.99) | 0.021 | 2.2×10⁻² | 49 | IVW | 0.045 | 65.829 | OR |
| CKD | East Asian | 1.31 (1.05; 1.63) | 0.112 | 1.5×10⁻² | 37 | MR Egger | 0.019 | 54.530 | OR |
|  | European | 0.93 (0.90; 0.97) | 0.020 | 4.7×10⁻⁴ | 51 | IVW | 0.448 | 50.634 | OR |
| COPD | East Asian | 0.97 (0.86; 1.08) | 0.058 | 5.6×10⁻¹ | 44 | MR Egger | 0.1 | 54.087 | OR |
|  | European | 0.99 (0.95; 1.04) | 0.023 | 6.5×10⁻¹ | 51 | MR Egger | 0.019 | 71.684 | OR |
| CRP | East Asian | -0.02 (-0.04; 0.01) | 0.014 | 1.8×10⁻¹ | 36 | IVW | 0.541 | 33.488 | mg/dL |
|  | European | 0.03 (0.00; 0.05) | 0.011 | 2.3×10⁻² | 44 | IVW | 0.253 | 48.759 | log(mg/L) |
| DBP | East Asian | -0.03 (-0.05; -0.01) | 0.011 | 7.8×10⁻³ | 37 | IVW | 0.11 | 46.639 | mmHg |
|  | European | -0.12 (-0.20; -0.03) | 0.044 | 7.1×10⁻³ | 45 | IVW | 0.079 | 57.798 | mmHg |
| Glaucoma | East Asian | 1.01 (0.90; 1.12) | 0.055 | 8.9×10⁻¹ | 36 | IVW | 0.125 | 44.767 | OR |
|  | European | 1.05 (1.01; 1.09) | 0.020 | 2.1×10⁻² | 51 | IVW | <0.001 | 91.251 | OR |
| Glucose | East Asian | -0.04 (-0.07; -0.02) | 0.012 | 5.8×10⁻⁴ | 38 | MR Egger | 0.033 | 53.122 | mg/dL |
|  | European | -0.03 (-0.04; -0.02) | 0.006 | 1.1×10⁻⁶ | 51 | MR Egger | 0.064 | 64.859 | mmol/L |
| HbA1c | East Asian | -0.01 (-0.04; 0.02) | 0.014 | 4.3×10⁻¹ | 36 | IVW | 0.101 | 45.987 | % |
|  | European | -0.00 (-0.01; 0.01) | 0.004 | 7.3×10⁻¹ | 51 | MR Egger | 0.862 | 38.405 | % |
| HF | East Asian | 0.85 (0.78; 0.94) | 0.049 | 1.3×10⁻³ | 36 | IVW | 0.061 | 48.791 | OR |
|  | European | 0.89 (0.86; 0.93) | 0.020 | 6.7×10⁻⁹ | 49 | IVW | 0.07 | 63.172 | OR |
| Intracerebral hemorrhage | East Asian | 0.69 (0.55; 0.87) | 0.115 | 1.5×10⁻³ | 34 | IVW | 0.003 | 59.529 | OR |
|  | European | 0.75 (0.66; 0.85) | 0.065 | 5.8×10⁻⁶ | 56 | IVW | 0.003 | 88.490 | OR |
| Ischemic stroke | East Asian | 1.04 (0.96; 1.12) | 0.041 | 3.5×10⁻¹ | 34 | MR Egger | 0.046 | 46.633 | OR |
|  | European | 0.96 (0.93; 0.99) | 0.014 | 2.8×10⁻³ | 34 | IVW | 0.28 | 37.239 | OR |
| LDL-C | East Asian | -0.04 (-0.09; 0.00) | 0.023 | 6.0×10⁻² | 34 | MR Egger | <0.001 | 61.296 | mg/dL |
|  | European | -0.15 (-0.16; -0.14) | 0.005 | 1.0×10⁻¹⁰⁰ | 58 | MR Egger | <0.001 | 133.351 | SD |
| Lp[a] | East Asian | -0.25 (-0.44; -0.07) | 0.095 | 7.7×10⁻³ | 38 | MR Egger | 0.029 | 53.685 | nmol/l |
|  | European | -2.70 (-3.27; -2.13) | 0.290 | 1.0×10⁻¹⁰⁰ | 60 | MR Egger | 0.023 | 81.429 | nmol/l |
| PAD | East Asian | 0.70 (0.60; 0.82) | 0.078 | 5.4×10⁻⁶ | 35 | IVW | 0.121 | 43.817 | OR |
|  | European | 0.96 (0.87; 1.05) | 0.046 | 3.5×10⁻¹ | 59 | MR Egger | <0.001 | 111.073 | OR |
| Pneumonia | East Asian | 0.89 (0.81; 0.99) | 0.050 | 2.6×10⁻² | 36 | IVW | 0.954 | 22.213 | OR |
|  | European | 0.87 (0.84; 0.90) | 0.018 | 6.7×10⁻¹⁶ | 58 | IVW | 0.026 | 79.517 | OR |
| **Outcome** | **Ancestry** | **MR estimate** | **se** | **p-value** | **n variants** | **model** | **Heterogeneity p-value** | **Q-statistic** | **unit** |
| PP | East Asian | 0.02 (-0.00; 0.05) | 0.013 | 9.0×10⁻² | 35 | MR Egger | 0.555 | 31.243 | mmHg |
|  | European | -0.17 (-0.26; -0.08) | 0.046 | 2.9×10⁻⁴ | 46 | IVW | <0.001 | 94.952 | mmHg |
| SBP | East Asian | 0.01 (-0.01; 0.04) | 0.013 | 4.0×10⁻¹ | 36 | MR Egger | 0.727 | 28.661 | mmHg |
|  | European | -0.40 (-0.57; -0.24) | 0.085 | 1.9×10⁻⁶ | 46 | MR Egger | 0.025 | 64.114 | mmHg |
| Subarachnoid hemorrhage | East Asian | 0.84 (0.66; 1.08) | 0.123 | 1.7×10⁻¹ | 35 | IVW | 0.024 | 52.140 | OR |
|  | European | 0.80 (0.71; 0.90) | 0.063 | 4.0×10⁻⁴ | 29 | IVW | <0.001 | 57.829 | OR |
| T2DM | East Asian | 0.99 (0.92; 1.05) | 0.032 | 6.5×10⁻¹ | 33 | IVW | 0.17 | 39.477 | OR |
|  | European | 1.00 (0.95; 1.06) | 0.027 | 9.2×10⁻¹ | 50 | MR Egger | 0.111 | 60.206 | OR |
| TG | East Asian | -0.12 (-0.14; -0.09) | 0.012 | 1.0×10⁻¹⁰⁰ | 37 | IVW | 0.378 | 38.012 | mg/dL |
|  | European | -0.12 (-0.13; -0.12) | 0.002 | 1.0×10⁻¹⁰⁰ | 68 | IVW | 0.002 | 105.807 | SD |
| Ventricular arrhythmia | East Asian | 1.09 (0.82; 1.45) | 0.145 | 5.3×10⁻¹ | 38 | MR Egger | 0.08 | 48.502 | OR |
|  | European | 0.91 (0.78; 1.06) | 0.078 | 2.0×10⁻¹ | 55 | IVW | 0.16 | 64.283 | OR |

Abbreviations**:** Apo-A1: apolipoprotein A, Apo-B: apolipoprotein B, Lp[a] lipoprotein a, CHD: coronary heart disease, CKD: chronic kidney disease, COPD: chronic obstructive pulmonary disease, CRP: C-reactive protein, DBP: diastolic blood pressure, Lp[a]: lipoprotein a, PAD: peripheral artery disease, PP: pulse pressure, SBP: systolic blood pressure, T2D: type 2 diabetes, TG: triglycerides.

| **Outcome** | **Unit** | **Interaction effect** | **Interaction se** | **Interaction p-value** |
| --- | --- | --- | --- | --- |
| Angina | OR | 0.952 (0.872;1.040) | 0.045 | 2.74×10⁻¹ |
| Any stroke | OR | 1.011 (0.917;1.113) | 0.049 | 8.23×10⁻¹ |
| Apo-A1 | g/l | -0.468 (-0.597;-0.339) | 0.066 | 1.02×10⁻¹² |
| Apo-B | g/l | 0.134 (-0.122;0.389) | 0.130 | 3.06×10⁻¹ |
| Asthma | OR | 0.754 (0.690;0.823) | 0.045 | 3.30×10⁻¹⁰ |
| CHD | OR | 1.074 (0.999;1.155) | 0.037 | 5.25×10⁻² |
| CKD | OR | 0.712 (0.569;0.889) | 0.114 | 2.81×10⁻³ |
| COPD | OR | 1.023 (0.906;1.156) | 0.062 | 7.05×10⁻¹ |
| CRP | mg/dL | 0.044 (0.009;0.079) | 0.018 | 1.31×10⁻² |
| DBP | mmHg | -0.087 (-0.175;0.002) | 0.045 | 5.42×10⁻² |
| Glaucoma | OR | 1.039 (0.927;1.165) | 0.058 | 5.12×10⁻¹ |
| Glucose | mg/dL | 0.015 (-0.011;0.042) | 0.014 | 2.55×10⁻¹ |
| HbA1c | % | 0.009 (-0.019;0.038) | 0.015 | 5.02×10⁻¹ |
| HF | OR | 1.040 (0.938;1.543) | 0.053 | 4.46×10⁻¹ |
| Intracerebral haemorrhage | OR | 1.074 (0.829;1.391) | 0.132 | 5.90×10⁻¹ |
| Ischemic stroke | OR | 0.923 (0.848;1.004) | 0.043 | 6.24×10⁻² |
| LDL-C | mg/dL | -0.107 (-0.153;-0.060) | 0.024 | 6.89×10⁻⁶ |
| Lp[a] | nmol/l | -2.444 (-3.043;-1.845) | 0.305 | 1.22×10⁻¹⁵ |
| PAD | OR | 1.367 (1.144;1.634) | 0.091 | 5.76×10⁻⁴ |
| Pneumonia | OR | 0.967 (0.872;1.073) | 0.053 | 5.31×10⁻¹ |
| PP | mmHg | -0.189 (-0.281;-0.095) | 0.048 | 7.78×10⁻⁵ |
| SBP | mmHg | -0.414 (-0.581;-0.246) | 0.086 | 1.33×10⁻⁶ |
| Subarachnoid haemorrhage | OR | 0.946 (0.721;1.242) | 0.139 | 6.90×10⁻¹ |
| T2DM | OR | 1.017 (0.937;1.104) | 0.042 | 6.79×10⁻¹ |
| TG | mg/dL | -0.009 (-0.033;0.015) | 0.012 | 4.66×10⁻¹ |
| Ventricular arrhythmia | OR | 0.827 (0.599;1.142) | 0.165 | 2.49×10⁻¹ |

Abbreviations**:** Apo-A1: apolipoprotein A, Apo-B: apolipoprotein B, Lp[a] lipoprotein a, CHD: coronary heart disease, CKD: chronic kidney disease, COPD: chronic obstructive pulmonary disease, CRP: C-reactive protein, DBP: diastolic blood pressure, Lp[a]: lipoprotein a, PAD: peripheral artery disease, PP: pulse pressure, SBP: systolic blood pressure, T2D: type 2 diabetes, TG: triglycerides.

**Supplemental Table 3. Interaction test of CETP Mendelian randomisation effect estimates on biomarkers and clinical outcomes between the East Asian and European population.**

**Supplemental Table 4. Baseline characteristics of biomarkers across European (UKB), East Asian (this study) and Chinese populations (Millwood *et al.*)**.

| **Biomarker** | **Unit** | **European** | **East Asian** | **Millwood *et al.*** | **Source** |
| --- | --- | --- | --- | --- | --- |
| HDL-C | mg/dl | 55.97 | 54.80 | 47.70 | <https://jamanetwork.com/journals/jamacardiology/fullarticle/2661813> |
| LDL-C | mg/dl | 137.03 | 130.90 | 91.40 | <https://jamanetwork.com/journals/jamacardiology/fullarticle/2661813> |
| TG | mg/dl | 153.99 | 133.00 | 139.80 | <https://jamanetwork.com/journals/jamacardiology/fullarticle/2661813> |
| Apo-A1 | g/l | 1.53 | 1.55 | 1.34 | [https://jamanetwork.com/journals/jamacardiology/fullarticle/2661816 https://www.ncbi.nlm.nih.gov/pmc/articles/PMC8792153/](https://jamanetwork.com/journals/jamacardiology/fullarticle/2661816https:/www.ncbi.nlm.nih.gov/pmc/articles/PMC8792153/) |
| Apo-B | g/l | 0.83 | 0.99 | 0.83 | [https://jamanetwork.com/journals/jamacardiology/fullarticle/2661816 https://www.ncbi.nlm.nih.gov/pmc/articles/PMC8792153/](https://jamanetwork.com/journals/jamacardiology/fullarticle/2661816https:/www.ncbi.nlm.nih.gov/pmc/articles/PMC8792153/) |
| Lp[a] | nmol/l | 44.63 | 32.90 | 29.70 | [https://jamanetwork.com/journals/jamacardiology/fullarticle/2661816 https://www.ncbi.nlm.nih.gov/pmc/articles/PMC8792153/](https://jamanetwork.com/journals/jamacardiology/fullarticle/2661816https:/www.ncbi.nlm.nih.gov/pmc/articles/PMC8792153/) |
| SBP | mmHg | 138.00 | 140.00 | 131.00 | <https://www.ncbi.nlm.nih.gov/pmc/articles/PMC3235021/table/dyr120-T2/?report=objectonly> |
| DBP | mmHg | 81.81 | 82.00 | 78.00 | <https://www.ncbi.nlm.nih.gov/pmc/articles/PMC3235021/table/dyr120-T2/?report=objectonly> |
| Glucose | mg/dl | 92.16 | 118.70 | 106.80 | <https://www.ncbi.nlm.nih.gov/pmc/articles/PMC3235021/table/dyr120-T2/?report=objectonly> |

Abbreviations: TG: triglycerides, Apo-A1: apolipoprotein A, Apo-B: apolipoprotein B, Lp[a] lipoprotein a, SBP: systolic blood pressure, DBP: diastolic blood pressure.

| **Outcome** | **Unit** | **East Asian  effect (95% CI)** | **East Asian se** | **East Asian**  **p-value** | **East Asian case/No. subjects** | **Millwood *et al.* effect (95% CI)** | **Millwood *et al*. se** | **Millwood**  ***et al.***  **p-value** | **Millwood *et al.* cases/No subjects** | **Interaction  (95% CI)** | **Interaction se** | **Interaction p-value** |
| --- | --- | --- | --- | --- | --- | --- | --- | --- | --- | --- | --- | --- |
| Apo-A1 | g/l | 0.67 (0.54;0.80) | 0.07 | 1.0×10⁻¹⁰ | 0/2325 | 1.29 (1.04;1.35) | 0.08 | 4.9×10⁻⁶⁵ | 0/17761 | -0.62 (-0.81;-0.42) | 0.10 | 5.6×10⁻¹⁰ |
| Apo-B | g/l | -0.18 (-0.44;0.07) | 0.13 | 1.6×10⁻¹ | 0/2553 | -0.27 (-1.69;1.15) | 0.72 | 7.1×10⁻¹ | 0/17761 | 0.90 (0.72;1.07) | 0.09 | 9.0×10⁻¹ |
| Asthma | OR | 1.26 (1.16;1.36) | 0.04 | 5.8×10⁻⁹ | 13015/162933 | 0.96 (0.49;1.88) | 0.38 | 9.0×10⁻¹ | 180/134996 | 0.30 (-0.38;0.98) | 0.35 | 3.9×10⁻¹ |
| CHD | OR | 0.89 (0.84;0.94) | 0.03 | 1.2×10⁻⁴ | 32512/146214 | 1.08 (0.95;1.22) | 0.06 | 2.2×10⁻¹ | 5767/119644 | -0.19 (-0.30;-0.07) | 0.06 | 4.9×10⁻¹ |
| CKD | OR | 1.31 (1.05;1.63) | 0.11 | 1.5×10⁻² | 2117/174345 | 1.06 (0.82;1.36) | 0.55 | 6.7×10⁻¹ | 1221/131596 | 0.25 (-0.84,1.34) | 0.56 | 6.5×10⁻¹ |
| COPD | OR | 0.97 (0.86;1.08) | 0.06 | 5.6×10⁻¹ | 19044/310689 | 1.01 (0.90;1.13) | 0.05 | 9.1×10⁻¹ | 6900/128506 | 0.96 (0.82;1.11) | 0.07 | 6.0×10⁻¹ |
| Glucose | mg/dl | -0.04 (-0.07;-0.02) | 0.01 | 5.8×10⁻⁴ | 0/133336 | -0.59 (-1.56;0.38) | 0.49 | 2.4×10⁻¹ | 0/149524 | 0.55 (-0.41;1.51) | 0.49 | 2.6×10⁻¹ |
| IS | OR | 1.04 (0.96;1.12) | 0.04 | 3.5×10⁻¹ | 22664/152022 | 0.94 (0.86;1.02) | 0.04 | 1.3×10⁻¹ | 13759/119644 | 0.10 (0.02;0.19) | 0.05 | 8.0×10⁻¹ |
| LDL-C | mg/dl | -0.04 (-0.09;0.00) | 0.02 | 6.0×10⁻² | 0/72866 | 0.24 (0.16;0.31) | 0.04 | 1.0×10⁻² | 0/17761 | -0.28 (-0.35;-0.20) | 0.04 | 3.8×10⁻¹⁰ |
| Lp[a] | nmol/l | -0.25 (-0.44;-0.07) | 0.10 | 7.7×10⁻³ | 0/2275 | -0.09 (-0.18;-0.00) | 0.04 | 4.0×10⁻² | 0/17761 | -0.16 (-0.35;0.03) | 0.1 | 1.2×10⁻¹ |
| Pneumonia | OR | 0.89 (0.81;0.99) | 0.05 | 2.6×10⁻² | 7423/171303 | 1.11 (0.92;1.33) | 0.18 | 2.7×10⁻¹ | 2446/132290 | -0.22 (-0.57;0.13) | 0.18 | 2.3×10⁻¹ |
| SBP | mmHg | 0.01 (-0.01; 0.04) | 0.01 | 4.0×10⁻¹ | 0/145505 | -0.73 (-0.24,-1.21) | 0.25 | 4.0×10⁻³ | 0/151047 | -0.74 (-0.25;1.23) | 0.25 | 3.1×10⁻³ |
| T2DM | OR | 0.99 (0.92; 1.05) | 0.03 | 6.5×10⁻¹ | 45383/132032 | 1.01 (0.90;1.12) | 0.05 | 9.2×10⁻¹ | 6972/125790 | -0.01 (-1.7;1.15) | 0.59 | 8.6×10⁻¹ |
| TG | mg/dl | -0.12 (-0.14; -0.09) | 0.01 | 1.0×10⁻¹⁰⁰ | 0/111667 | -1.46 (2-2.4;-0.51) | 0.48 | 2.4×10⁻³ | 0/17761 | 1.34 (0.39;2.28) | 0.48 | 5.2×10⁻³ |

**Supplemental Table 5. Comparison of CETP effects on clinical outcomes and biomarkers with Millwood *et al.* in the East Asian population.**

Abbreviations**:** Apo-A1: apolipoprotein A, Apo-B: apolipoprotein B, CHD: coronary heart disease, CKD: chronic kidney disease, COPD: chronic obstructive pulmonary disease, IS: ischemic stroke, Lp[a] lipoprotein a, Lp[a]: lipoprotein a, SBP: systolic blood pressure, T2D: type 2 diabetes, TG: triglycerides.
